# Safety, immunogenicity, and relative efficacy of a parenteral trivalent rotavirus subunit vaccine candidate (TV P2-VP8) in healthy Ghanaian, Malawian, and Zambian infants

**DOI:** 10.1101/2025.09.15.25335434

**Authors:** Tushar Tewari, George Armah, Nigel A. Cunliffe, Caroline C. Chisenga, Desiree Witte, Vida Kukula, Khuzwayo C. Jere, Michelo Simuyandi, Susan Damanka, Edson Mwinjiwa, Katayi Kazimbaya, Frank Atuguba, John Williams, Roma Chilengi, Joanne Csedrik, Chris Gast, Alan Fix, Stan Cryz

## Abstract

**Background:** While live oral rotavirus vaccines (LORVs) are effective at preventing severe rotavirus gastroenteritis (SRVGE), they perform sub-optimally in the settings with the highest burden. Parenteral rotavirus vaccines could have greater efficacy by bypassing the gut, where interference with LORVs may occur. A novel parenteral rotavirus subunit vaccine (TV P2-VP8) was evaluated in a Phase 3 study for safety, immunogenicity, and relative efficacy in healthy African infants.

**Methods:** Participating infants were randomized at 6–8 weeks of age to receive either three doses of TV P2-VP8 intramuscularly (IM) plus oral placebo or two doses of ROTARIX® (oral) plus IM placebo. The trial was designed to enroll a total of 8,200 participants in two stages separated by an interim analysis conducted upon accrual of a predetermined number of SRVGE cases. Data at interim analysis indicated a low probability of demonstrating TV P2-VP8 superiority compared to ROTARIX in prevention of SRVGE, and the study was closed early.

**Results:** 4,055 infants were enrolled and provided safety data. TV P2-VP8 was safe and well tolerated, with no safety concerns indicated. Virus neutralizing and IgG binding antibody seroresponses to TV-P2-VP8 were detected in most participants. The crude attack rates of SRVGE in the per-protocol population (n=3,477) were 4.51% (TV P2-VP8) and 2.57% (ROTARIX). The relative vaccine efficacy (RVE) against SRVGE for TV P2-VP8 was - 77.68% (95% CI, -162.38, -21.54). The RVE against SRVGE in the first and second year of life was -91.67% (95% CI, -199.20, -24.35) and -31.97% (95% CI, -220.37, 44.49), respectively. The RVE against rotavirus gastroenteritis of any severity was - 47.78% (95% CI, -89.63, -15.59).

**Conclusions:** TV P2-VP8 provided inferior protection against SRVGE compared to ROTARIX. The vaccine candidate was safe, immunogenic, and well-tolerated by infants. Continued efforts are needed to identify alternative rotavirus vaccines with improved efficacy in LMICs.

**Highlights:**

- The TV P2-VP8 vaccine candidate was safe and immunogenic.
- TV P2-VP8 was inferior to ROTARIX® in preventing severe rotavirus gastroenteritis.
- Study provides insight to guide future development of rotavirus vaccine candidates.

## 1. Introduction

Pediatric diarrheal disease burden has significantly declined over the last three decades due to improved implementation of prevention and control strategies. However, diarrhea remains a leading cause of morbidity and mortality in children younger than 5 years of age, predominantly in low- and middle-income countries (LMICs). Despite the availability of rotavirus vaccines, rotavirus remains a principal cause of severe diarrheal disease globally, accounting for an estimated 176,000 deaths in 2021 [1]. The World Health Organization (WHO) recommends live, oral rotavirus vaccines (LORVs) for all infants worldwide. A large portion of the world still has no access to rotavirus vaccines as global vaccination coverage is estimated at 59% [2][3]. LORVs are highly effective (∼90%) in high-income settings (HICs) [4][5]; however for reasons not well understood, these vaccines do not perform optimally in LMIC populations, where the efficacy is modest (∼50%) [6][7][8][9] and the burden is highest, illustrating the need for improved vaccines.

The discrepancy in vaccine performance between LMICs and HICs is not a unique feature of LORVs. It has also been observed with other oral, live-attenuated enteric vaccines, such as those for cholera and poliovirus. Some theories explaining reduced vaccine efficacy in resource-limited regions include environmental enteropathy, interference by co-infection with other enteric pathogens, genetic predispositions and relatively higher levels of maternally-derived antibodies because of repeated exposure. Known factors like oral polio vaccine interference on LORV responses support the theory of pathogen interference[10][11].

One potential strategy to improve the efficacy of rotavirus vaccination is the development of inactivated or subunit, parenterally administered vaccines. Unlike oral vaccines, parenteral vaccines bypass many of the barriers associated with the gut environment. This approach is supported by the success of injectable vaccines to prevent other major enteric diseases, including those caused by poliovirus and *Salmonella* Typhi. These examples demonstrate the potential of parenteral vaccines to overcome mucosal interference and elicit robust systemic immunity, making them a compelling alternative approach for rotavirus prevention.

The parenterally administered non-replicating trivalent P2-VP8 subunit rotavirus vaccine (TV P2-VP8) demonstrated promise in early-phase clinical trials [12][13][14]. The vaccine was safe and well-tolerated in South African infants when concomitantly administered with recommended Expanded Programme on Immunization (EPI) vaccines. Immunization engendered a robust serum neutralizing antibody response and significantly reduced fecal shedding from a subsequent “challenge” with oral ROTARIX vaccine strain compared to a placebo group, thus indicating a potentially protective local gut effect [13][14]. Based upon these promising results, a multi-country Phase 3 study was conducted to evaluate the safety, relative vaccine efficacy (RVE), and immunogenicity of the TV P2-VP8 vaccine candidate in prevention of severe rotavirus gastroenteritis (SRVGE) using ROTARIX as a comparator vaccine.

## 2. Methods

### 2.1 Ethical Considerations

The study was conducted at the Dodowa Health Research Centre, Ghana; Malawi Liverpool Wellcome Research Programme (MLW), Malawi; and the Centre for Infectious Disease Research in Zambia (CIDRZ). The study protocol was reviewed and approved by the Institutional Review Board (IRB)/ Independent Ethics Committee (IEC) from study sites and the Western Institutional Review Board. All participants were enrolled after written consent was obtained from their parents or legal guardian. The study was carried out in compliance with the Good Clinical Practice (GCP) guidelines, as described in the ICH Harmonized Tripartite Guideline for GCP (E6) 2016, Declaration of Helsinki, concerning medical research in humans (Revised Brazil, 2013) and all local requirements and guidelines.

### 2.2 Study Design

This Phase 3 study was a randomized, double-blind, double-dummy, endpoint driven, group-sequential study. The primary endpoint was the incidence of SRVGE, defined as laboratory-confirmed RVGE, with a Vesikari score ≥11 [15]. The study was designed to evaluate superiority of TV P2-VP8 to a licensed comparator rotavirus vaccine (ROTARIX) for the prevention of SRVGE. The participating infants were randomized 1:1 to receive either three doses of intramuscular (IM) TV P2-VP8 plus two doses of oral placebo (oral rehydration solution; ORS), or oral two doses of ROTARIX plus three doses of IM placebo (normal saline). Both placebo formulations were needed to maintain the blind. The study vaccine was administered concomitantly with EPI vaccines mandated by the local EPI program, at a separate injection site. Participants received three doses of TV P2-VP8 or two doses of ROTARIX with corresponding placebos at monthly intervals starting at 6–8 weeks of age, followed by active surveillance for gastroenteritis (GE) (Supplementary Table 1). The event-driven trial was designed to enroll participants in two stages separated by an interim analysis, conducted when at least 30 SRVGE cases were accrued with progression to the second stage dependent on meeting pre-specified criteria. An independent Data and Safety Monitoring Board (DSMB) was established to review the unblinded SRVGE case split against predefined criteria. The criteria were set based upon the probability of demonstrating superiority. Following the review of the interim analysis data, the DSMB would recommend whether the study should continue to full enrollment, or the study should be closed early.

### 2.3 Study Participants

Participants were 6- to 8-week-old healthy infants whose parents or legal guardians gave written informed consent and who were residents of the study area. Eligibility was assessed through medical history and physical examination, and investigators used their clinical judgment in ascertaining participant’s eligibility for inclusion in the study. Key exclusion criteria were the presence of severe malnutrition or any systemic disorder that would compromise the participant’s health or result in non-compliance to the protocol; history of premature birth (<37 weeks gestation) and/or birth weight of <2.5 kg; history of congenital abdominal disorders, intussusception, or abdominal surgery; prior receipt of rotavirus vaccine; known sensitivity or allergy to any components of the study vaccine; immunoglobulin therapy or chronic immunosuppressant medications; and contraindication to any EPI vaccine.

### 2.4 Active surveillance of gastroenteritis

Active GE surveillance was conducted through weekly home-visit and phone contact. Whenever a participant experienced a GE episode (≥3 looser than normal stools within 24 hours), parents/family members were to notify the study team and collect approximately 5 mL of stool in study provided stool containers. Parents were also provided with a diarrheal diary card to record axillary temperature, number and duration of vomiting episodes, number and duration of looser than normal stools, any treatment given (including oral and/or intravenous rehydration), and duration of hospital stay (if any) during the episode. The participants with GE episodes were to be evaluated by a qualified clinician to assess elements of dehydration and to determine appropriate clinical management during illness.

### 2.5 Randomization and Blinding

Participants were randomized at a ratio of 1:1 to study arms. Block-randomization, facilitated by a validated interactive web response system (IWRS), was stratified by site and with a block size of 6 to ensure balance within each site. After obtaining consent, a unique participant identification code (Participant ID) was allocated to each participant through IWRS, and eligible infants were randomized on the day of the first vaccination. At each site, an unblinded pharmacist prepared doses for each vaccine and accompanying placebo. After randomization, a blinded study nurse or physician administered the study vaccines.

### 2.6 Investigational Products

The TV P2-VP8 vaccine candidate (Lot number CTRAV401 and CTRAV402) is composed of recombinant, truncated VP8 rotavirus structural proteins representing the P[4], P[6], and P[8] serotypes derived from the DS-1 P[4], 1076 [P6], and Wa [P8] strains, fused to the P2 CD4+ T cell epitope of tetanus toxin. The purified proteins were then blended in equal amounts and co-formulated with aluminum hydroxide. Each 0.5 mL dose, administered intramuscularly, contains 30 μg of each of the three antigens, for a total of 90 μg of protein per dose. The vaccine candidate was manufactured by S. K. Biosciences (Seoul, South Korea).

ROTARIX is a WHO prequalified and licensed rotavirus vaccine which contains the attenuated RIX4414 strain of human rotavirus expressing the G1P[8] type and contains no less than 10^6.0^ CCID_50_ (cell culture infectious dose 50%) of the RIX4414 strain. Each dose is composed of 1.5 mL of the liquid vaccine, which is administered orally. The vaccine is manufactured by GSK Biologicals (Rixensart, Belgium). Separate lots were used in Ghana (AROLC602AA, AROLC982BC), Malawi (AROLDO11BA, AROLDOO5AA) and Zambia (AROLC816AA, AROLD113AA, AROLD150AA) depending on the availability in the respective EPI programs.

The oral placebo used in the TV P2-VP8 arm was a clear, colorless, locally licensed ORS, while the IM placebo used in the ROTARIX arm was composed of 0.9% Sodium Chloride for injection USP.

## 3. Outcome Assessment

### 3.1 Efficacy

The information from the diarrheal diary card was used to derive severity scores for each GE episode, according to the Vesikari Scoring System [15]. An episode of GE with a score of <7 was classified as ‘mild’, 7–10 as ‘moderate’, ≥11 as ‘severe’, and ≥15 as ‘very severe’. Stool samples were to be collected from all participants with a GE episode to determine the presence of rotavirus. Diagnosis of rotavirus infection was determined at the clinical site laboratory by enzyme immunoassay (EIA) using the Rotaclone^®^ rotavirus antigen detection following manufacturer’s instructions [16]. The stool samples from all sites for GE episodes with moderate and greater severity that were positive for rotavirus were sent to the WHO West Africa Rotavirus Regional Reference Laboratory at Noguchi Memorial Institute for Medical Research Accra, Ghana, for genotyping.

A case of SRVGE constituted a clinical endpoint for the primary analysis of efficacy. For participants experiencing more than one SRVGE episode, only the first episode was counted toward the primary endpoint. This was an event driven study in which the primary analysis was an assessment of efficacy against SRVGE occurring ≥14 days after the receipt of all scheduled study vaccinations in participants who were free from any major protocol deviations. The primary analysis was to occur when at least 99 cases of SRVGE were accrued. Interim analysis occurred after confirmation of ≥30 cases of SRVGE occurring at least two weeks after receipt of all study vaccinations. The interim analysis included a binding assessment to determine if the study should proceed to full enrollment.

### 3.2 Safety

All participants were monitored for 30 minutes after each vaccination for immediate adverse events (IAEs). A subset of participants (reactogenicity cohort, n=1,207) was provided with a post-immunization diary card to record solicited adverse events (AEs) (pain or tenderness, erythema/redness, swelling/induration, fever, diarrhea, irritability, decreased activity, decreased appetite, and vomiting) daily for 7 days after each vaccination. Solicited AEs were not assessed for causality and were considered related to study products by default. Unsolicited AEs were reported spontaneously by the participant’s parents, observed by the study personnel during study visits, or identified during review of medical records/source documents. The severity of all AEs / SAEs that occurred during the study were graded as per the guidance document by the Division of AIDS Table for Grading the Severity of Adult and Pediatric Adverse Events, version 2.0, July 2017 of the US National Institute of Health [17].

For non-serious, unsolicited AEs, only those of Grade 2 or higher and occurring within 28 days of vaccination were captured in the study database. Serious AEs were recorded and reported from the first study vaccination to the end of the study period for each study participant.

A Protocol Safety Review Team composed of physicians from within the study team and an independent physician met periodically to review blinded safety data. A DSMB composed of independent physicians and a biostatistician provided unblinded review of safety and efficacy data.

### 3.3 Immunogenicity

Blood samples were collected at baseline and 28 days after the third vaccination for all participants. Immunogenicity was evaluated in a subset of participants, which included all endpoint cases and vaccination-, age- and site-matched controls (no rotavirus GE) in a 1:3 (case: control) ratio. IgG and IgA binding antibodies to the 3 antigens contained in the TV P2-VP8 vaccine candidate were measured by ELISA and neutralizing antibody levels were determined as previously described [18]. Serum from participants immunized with ROTARIX were tested for levels of IgA antibody using a whole virus lysate enzyme-linked immunosorbent assay (ELISA) where the lysate was derived from a ROTARIX-like strain. The serum from participants that received the TV P2-VP8 vaccine candidate were not tested in this assay as it primarily detects anti-VP6 antibody and the parenteral vaccine lacks a VP6 component. Tests were performed at the Laboratory for Specialized Clinical Studies, Cincinnati Children’s Hospital Medical Centre (CCHMC), Cincinnati, Ohio, USA.

The IgG and IgA seroresponse rates were defined by ≥4-fold increase in antibody titers between baseline and 28 day post-third study injection. A significant neutralizing antibody increase was ≥2.7-fold increase over baseline. Both IgG and neutralizing antibody titers were adjusted for maternal antibody decay [13].

### 3.4 Statistical Considerations

For sample size calculations, the vaccine efficacy against SRVGE in the ROTARIX arm was assumed to be 50% and the attack rate of SRVGE through 2 years of age was assumed to be no less than 1.7%, an assumption drawn from a relevant prior study of an alternative LORV, as the lower confidence bound of the attack rate [8] Sample size was computed using exact group sequential methodology for binary data [19] and implemented in the gsDesign [20] package in R [21]. A total sample size of 8200 participants was required to achieve 90% power with a 1-sided type 1 error rate of 2.5% and considering a 5% drop-out rate. At primary analysis, the study was to sequentially examine a hypothesis of non-inferiority in the per-protocol efficacy population, followed by evaluation for superiority. A conclusion of non-inferiority (NI) was to be made if it was demonstrated that RVE > -30% and a conclusion of superiority was to be made if a RVE of > 0% was demonstrated, using the lower confidence bound of the two-sided 95% confidence interval for RVE, derived using the exact binomial method for TV P2-VP8 case count, conditional on the total, and adjusted for follow-up time in each group. Only the first case of SRVGE of a given type and the preceding follow-up time were used in incidence and RVE computations. A final analysis was to be performed after all cases were collected, following study closure.

RVE is expressed as the percent reduction in SRVGE attack rate (AR) in the TV P2-VP8 group relative to the active control (ROTARIX) group and computed as RVE = (1-RR)*100, where RR was the ratio of SRVGE AR in the TV P2-VP8 group relative to the active control (ROTARIX) group. The primary efficacy analysis was conducted in the per-protocol efficacy population (PP-EFF) and supported by a supplementary analysis in the intention-to-treat efficacy population (ITT-EFF). PP-EFF population included participants who received all vaccine doses per-protocol, had at least one follow-up contact/visit occurring 14 days after the third vaccine dose, and had no major protocol deviations considered likely to significantly interfere with the efficacy assessment. Participants randomized into the study who had at least one post-vaccination follow-up visit and were part of ITT-EFF population Secondary efficacy analyses included RVE against RVGE of any severity, against very severe RVGE (VSRVGE), against SRVGE in the first and second year of life, against SRVGE over the entire follow-up period, SRVGE associated with rotavirus serotypes included in the vaccine, and against SRVGE with hospitalization.

Safety analysis included analysis of immediate AEs, solicited AEs, unsolicited AEs, and SAEs. The primary variables for evaluation of the safety data were the percentage of participants with SAEs through 28 days after the last dose of study vaccine and AEs Grade ≥2 through 28 days after the last dose of study vaccine. Solicited AEs were summarized by site, severity and term, using the count and percentage of participants experiencing such an event. The primary method for safety data evaluations was descriptive without formal hypothesis testing. An overall summary presented the number and percentage (two-sided exact 95% CI) of AEs by MedDRA System Organ Class (SOC) and preferred term (PT), by group for all participants in the safety population. Exact CIs were constructed using the Clopper-Pearson method.

Among the group randomized to receive TV P2-VP8, the number and percentage of participants with antibody seroresponse were presented along with two-sided 95% CIs for IgG, IgA and serum neutralizing antibodies. For each assay and antigen, the geometric mean titer (GMT) and 95% CI was calculated as the antilog (base 10) of the estimated mean value obtained of the log_10_ antibody titer. The Geometric Mean Fold Rise (GMFR) along with two-sided 95% CIs for each group was calculated as for the GMT. Inverse-probability weighting was used to account for the case-control sampling design.

## 4. Results

### 4.1 Interim Analysis

An interim analysis was conducted after the accrual of 36 cases of SRVGE; 22 occurred in the TV P2-VP8 arm and 14 in the ROTARIX arm, which met the predetermined criterion to terminate the study. All participants who were age-eligible to receive LORV according to the local EPI program guidelines were unblinded, and those randomized to the TV P2-VP8 arm were offered ROTARIX. The DSMB recommended that all participants, including those unblinded and those remaining blinded, be followed up for an additional 6 months from the date of interim analysis for safety and GE surveillance, after which the study would close. A final efficacy analysis was performed, which included all participants with follow-up prior to unblinding for those eligible to receive crossover LORV and through the end of the study for those who were ineligible to receive catch up LORV and remained blinded.

### 4.2 Final Analysis

A total of 4,152 participants were enrolled in the study across three sites, of which a total of 4,055 participants received at least one dose of the study vaccine (2,031 received TV P2-VP8 and 2,024 received ROTARIX) and constituted the safety population. A total of 2,973 (73.3%) participants completed the study (2 years of age at final follow up), and 1,082 (26.7%) participants discontinued the study before completion (Fig 1). The most common reason for early termination was the COVID-19 pandemic as the study was suspended due to nationwide lockdown in Zambia. A total of 501 participants in the TV P2-VP8 arm were offered cross-over ROTARIX vaccination at sites in Zambia and Malawi following the interim analysis and decision to terminate the study early. None of the participants in Ghana received cross-over vaccination as they were over the eligible age to receive ROTARIX per their national EPI program. Overall, 1,728 (84.9%) participants in the TV P2-VP8 group and 1,749 (86%) participants in the ROTARIX group were included in the PP-EFF population. A total of 1,909 (93.8%) participants in the TV P2-VP8 group and 1,919 (94.4%) participants in the ROTARIX group were part of ITT-EFF population (Supplementary Fig 1).

**Figure 1:**
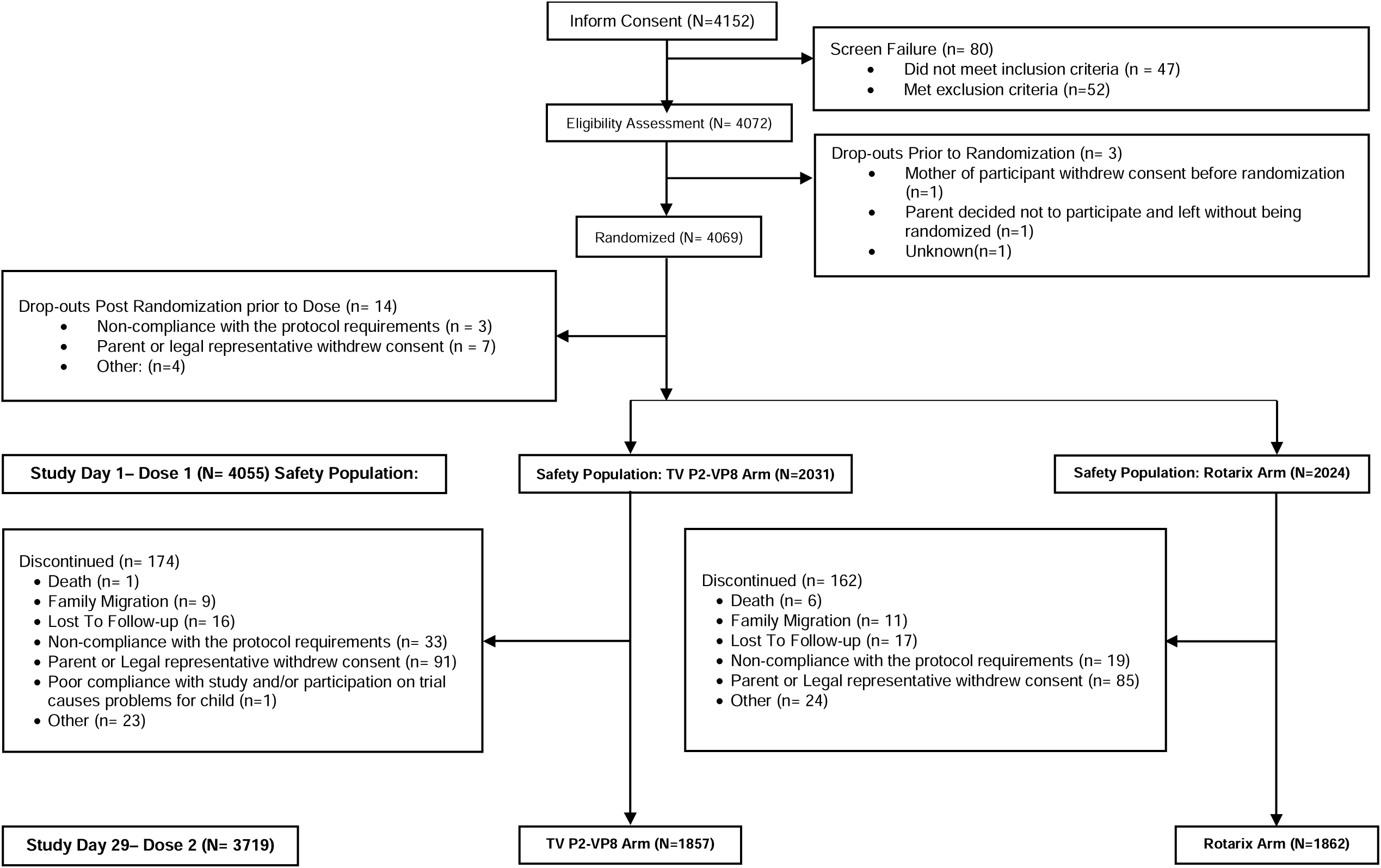
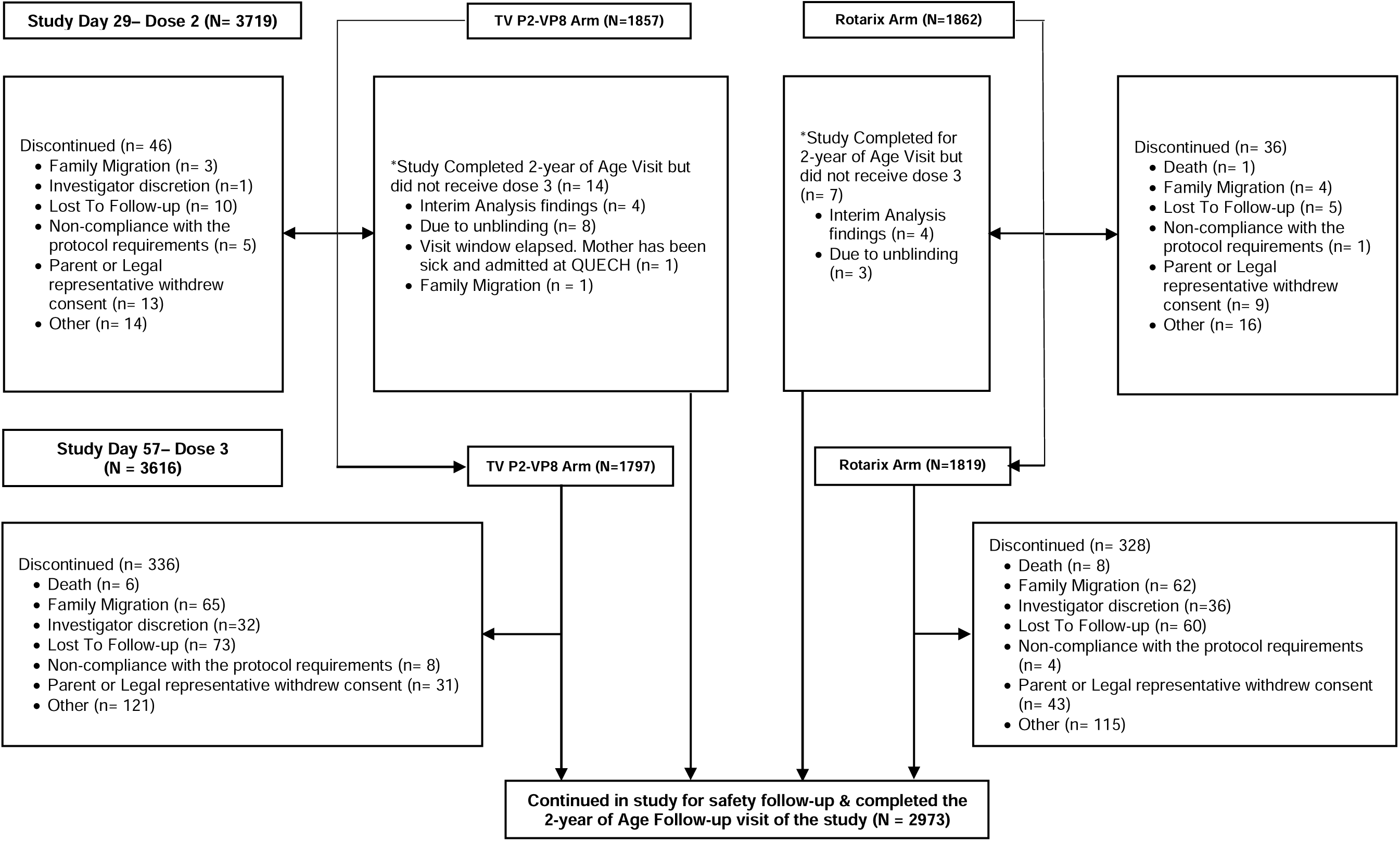
Study Flow.

Demographic characteristics were similar across study groups The proportion of male and female participants was comparable in both the TV P2-VP8 (51.4% and 48.6%) and ROTARIX (48.6% and 51.4%) group. Almost all the participants enrolled in the study were of Black African race (Table 1). Baseline characteristics with respect to mean age, weight, and length of the participants at enrollment were also comparable in both the study groups.

**Table 1:** Summary of Baseline Characteristics, Demographics and Other Characteristics – Safety Population.

| **Demographics Characteristic** | **Statistic** | **TV P2-VP8**<br>**(N=2031)** | **Rotarix**<br>**(N=2024)** |
| --- | --- | --- | --- |
| **Sex** |  |  |  |
| Male | n (%) | 1044 (51.4) | 984 (48.6) |
| **Race** |  |  |  |
| Black African | n (%) | 2030 (100.0) | 2022 (99.9) |
| Asian | n (%) | 1 (0.0) | 1 (0.0) |
| Other | n (%) | 0 | 1 (0.0) |
| **Age at Enrollment (Days)** | Mean (SD) | 44.9 (2.99) | 44.9 (3.01) |
|  | (Min, Max) | (42, 55) | (42, 55) |
| **Weight at Enrollment (Kg)** | Mean (SD) | 4.79 (0.610) | 4.77 (0.618) |
|  | (Min, Max) | (2.6, 7.0) | (3.0, 7.0) |
| **Length at Enrollment (cm)** | Mean (SD) | 54.30 (2.266) | 54.27 (2.245) |
|  | (Min, Max) | (45.0, 62.0) | (45.0, 63.5) |

#### 4.2.1 Safety Results

Safety was evaluated in all the participants who received at least one dose of study vaccine (n=4055). The participants who experienced at least one solicited AE following any vaccination (including IAEs) across both the groups were comparable (77.5% and 76.4% of participants in the TV P2-VP8 and ROTARIX groups, respectively) [Table 2; Figure 2]. The incidence of local solicited AEs with study injections were comparable in both groups [35.2% for TV P2-VP8 versus 34.1% for ROTARIX]. Additionally, all local solicited events were ≤ Grade 2 in intensity, with most participants (>90%) presenting with Grade 1 events. The most common local solicited event was injection-site pain or tenderness [290 events in 178 (29.4%) participants in TV P2-VP8 group and 273 events in 175 (29.1%) participants in the ROTARIX group following placebo injections]. The percentage of participants experiencing local solicited AEs decreased with subsequent dosing (Supplementary Table 2).

**Figure 2:**
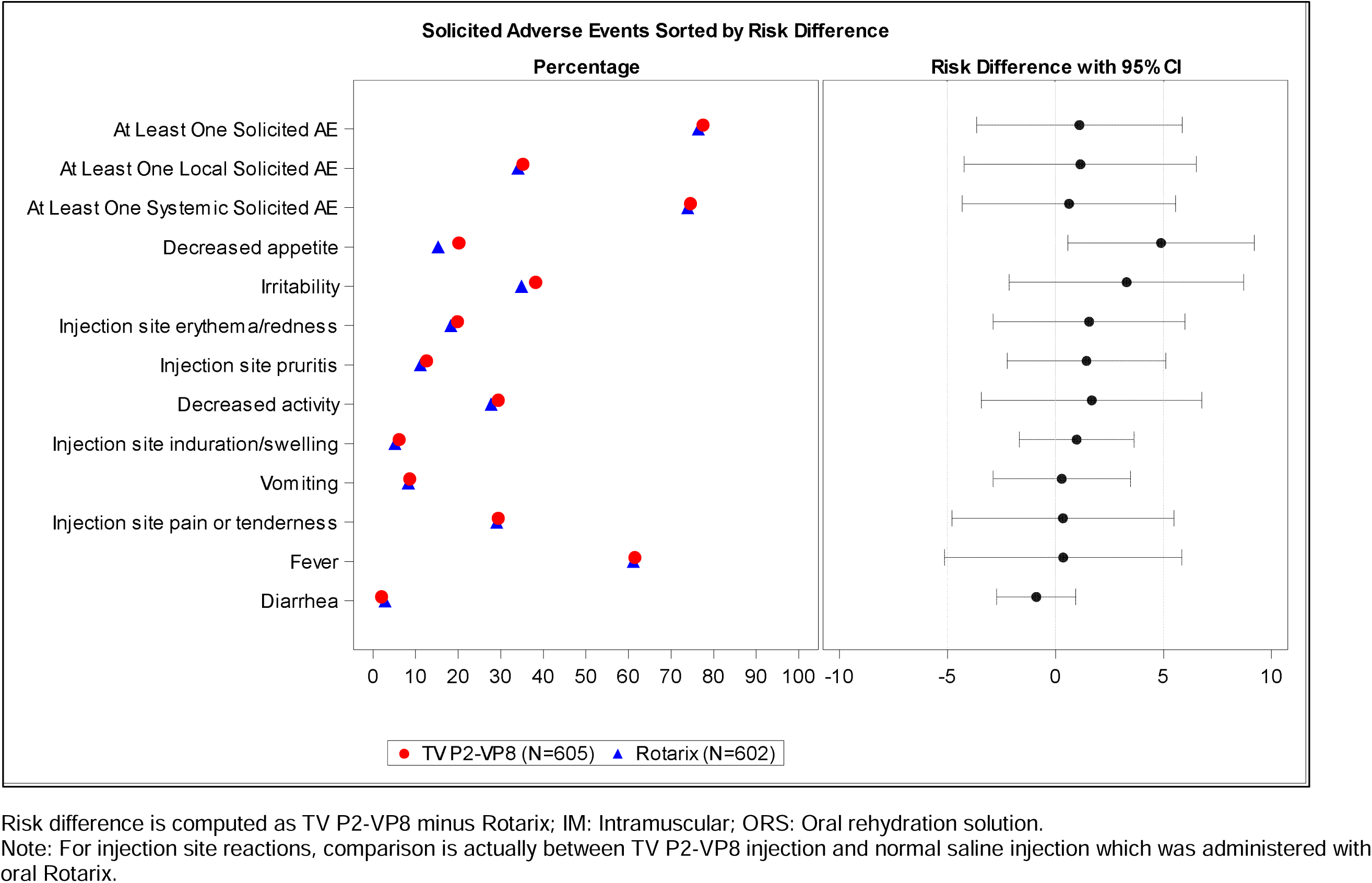
Risk Difference Plot to Compare Percentage of Solicited AEs in Two Treatment Arms, TV P2-VP8 (IM) plus ORS (oral) against Rotarix (oral) plus normal saline(IM) – Reactogenicity Population.

**Table 2:**
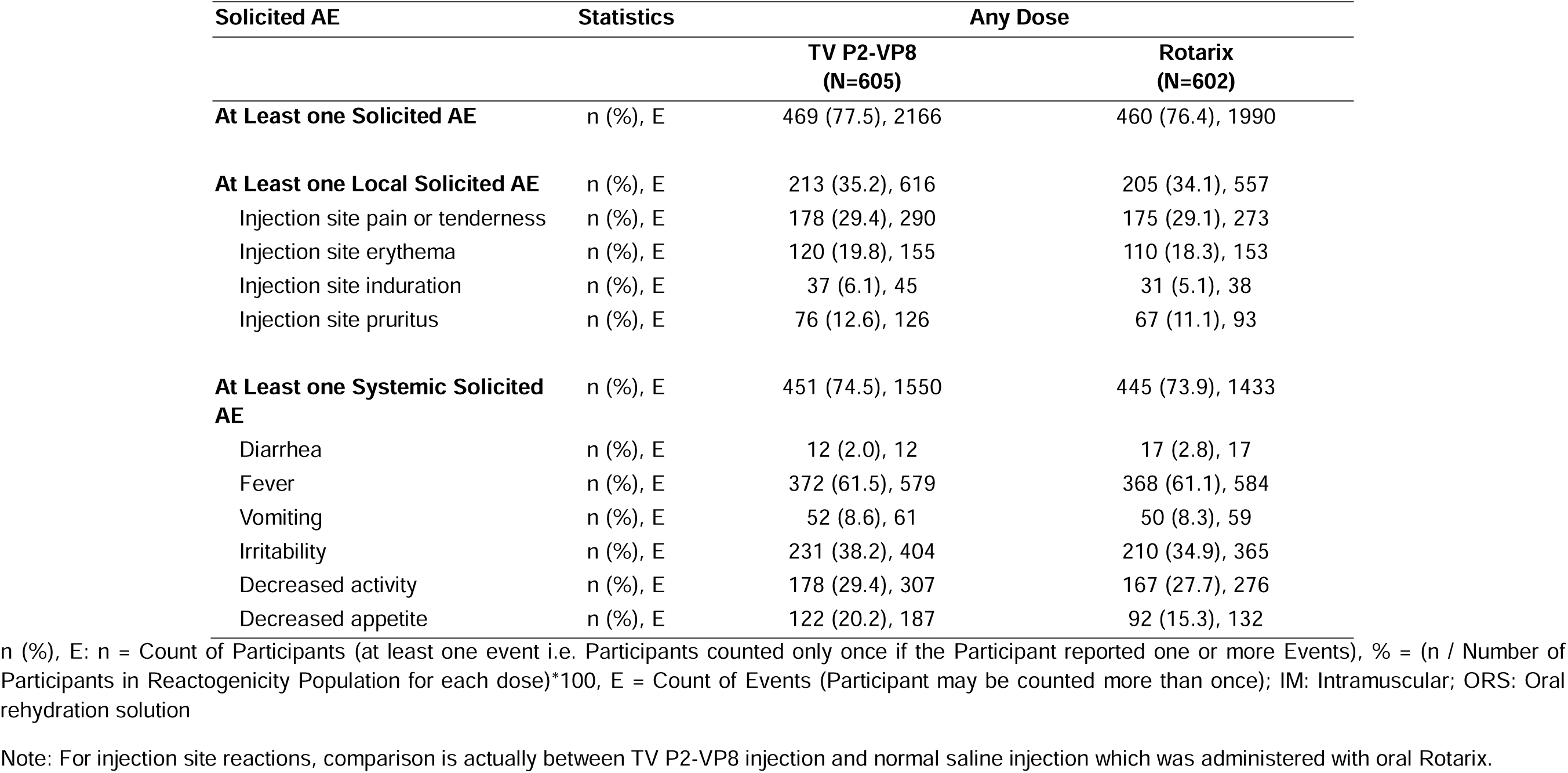
Overview of Reactogenicity during the 7 Days - TV P2-VP8 (IM) plus ORS (oral) against Rotarix (oral) plus normal saline (IM), Reactogenicity Population.

The incidence for all the systemic solicited events except decreased appetite, were comparable between the two groups [74.5% versus 73.9%, TV P2-VP8 and ROTARIX, respectively]. Participants in the TV P2-VP8 arm reported significantly higher incidence of decreased appetite. The majority of systemic solicited events were Grade 1 (60.2% versus 58.0%) or Grade 2 (11.7% versus 14.5%) in severity. The most common systemic solicited AE in both groups was fever, reported in approximately 60% of participants (Table 2).

Overall, the incidence of unsolicited AEs (of ≥ Grade 2 and reported within 28 days following vaccination) was comparable in both the groups (32.5% of participants in the TV P2-VP8 group and 30.5% of participants in the ROTARIX group; Table 3). The most frequently reported unsolicited AEs were upper respiratory tract infection and rhinitis. The majority of AEs were considered not related to the study vaccines, except for 18 events [12 events in the TV P2-VP8 and 6 events in the ROTARIX group]. Notably, most of these were reactogenicity events in the non-reactogenicity cohort, hence they were assessed for causality and were deemed related to the study vaccines. Most of the unsolicited AEs (>90%) reported were Grade 2 in severity.

**Table 3:** Summary of Unsolicited Adverse Events Through 28 Days After Last Dose: Overall – Safety Population.

| Category of Unsolicited AE | Statistics | Any Dose |  |
| --- | --- | --- | --- |
|  |  | TV P2-VP8<br>(N=2031) | Rotarix<br>(N=2024) |
| **At Least one Unsolicited AE** | n (%), E | 661 (32.5), 973 | 618 (30.5), 946 |
| **At Least one ≥ Grade 3 Unsolicited AE** | n (%), E | 32 (1.6), 34 | 29 (1.4), 30 |
| **At least one related Unsolicited AE** | n (%), E | 11 (0.5), 12 | 6 (0.3), 6 |
| **At least one SAE** | n (%), E | 32 (1.6), 34 | 28 (1.4), 29 |
| **Common Unsolicited AEs** | n (%), E |  |  |
| Upper respiratory tract infection | n (%), E | 414 (20.4), 535 | 425 (21.0), 558 |
|  | 95% CI | [18.65, 22.20] | [19.24, 22.84] |
| Rhinitis | n (%), E | 49 (2.4), 55 | 46 (2.3), 47 |
|  | 95% CI | [1.79, 3.18] | [1.67, 3.02] |
| Respiratory tract infection | n (%), E | 29 (1.4), 31 | 22 (1.1), 25 |
|  | 95% CI | [0.96, 2.04] | [0.68, 1.64] |

n (%), E: n = Count of Participants (at least one event i.e. Participants counted only once if the Participant reported one or more Events), % = (n / Number of Participants in Safety Population for whom specific safety endpoint data is available)\*100, E = Count of Events (Participant may be counted more than once)
Unsolicited AEs, except for SAEs, were only recorded in the database if grade ≥2 and occurring within 28 days of a vaccination
Unsolicited AEs also include IAEs & SAEs which are reported within 28 days post each dose.

A total of 307 SAEs were reported in the study — 148 SAEs were reported in the TV P2 VP8 group and 159 SAEs were reported in the ROTARIX group. Six events in TV P2 VP8 vaccine group and 16 events in ROTARIX group were fatal. All SAEs were assessed, and only one SAE of hyperthermia following vaccination with TV P2-VP8 vaccine was determined to be related to study vaccines. Two events of intussusception were reported following the third dose in the TV P2-VP8 group, which were considered not related to the study vaccine and recovered without sequelae. There was no significant difference observed in vital sign measurements and physical examinations between the two groups after any of the study vaccinations.

#### 4.2.2 Immunogenicity Results

Immunogenicity was evaluated in a case-control subset of participants which included all primary endpoint cases (n=108) and their matched controls (n=304). IgG seroresponse rates to TV P2-VP8 exceeded 97% for each vaccine P-type (Figure 3). Seroresponse rates for neutralizing antibodies in the TV P2-VP8 arm were 65%, 44%, and 63% for DS-1 (P[4]), 1076 (P[6]), and Wa (P[8]) strains, respectively (Figure 3). The IgA seroresponse rate among TV P2-VP8 recipients was ∼15% across all three serotypes. GMFRs paralleled those seen for IgG and neutralizing antibodies. Approximately 46% of ROTARIX recipients mounted a significant rise in IgA titer as determined in the whole virus lysate ELISA (data not shown).

**Figure 3.**
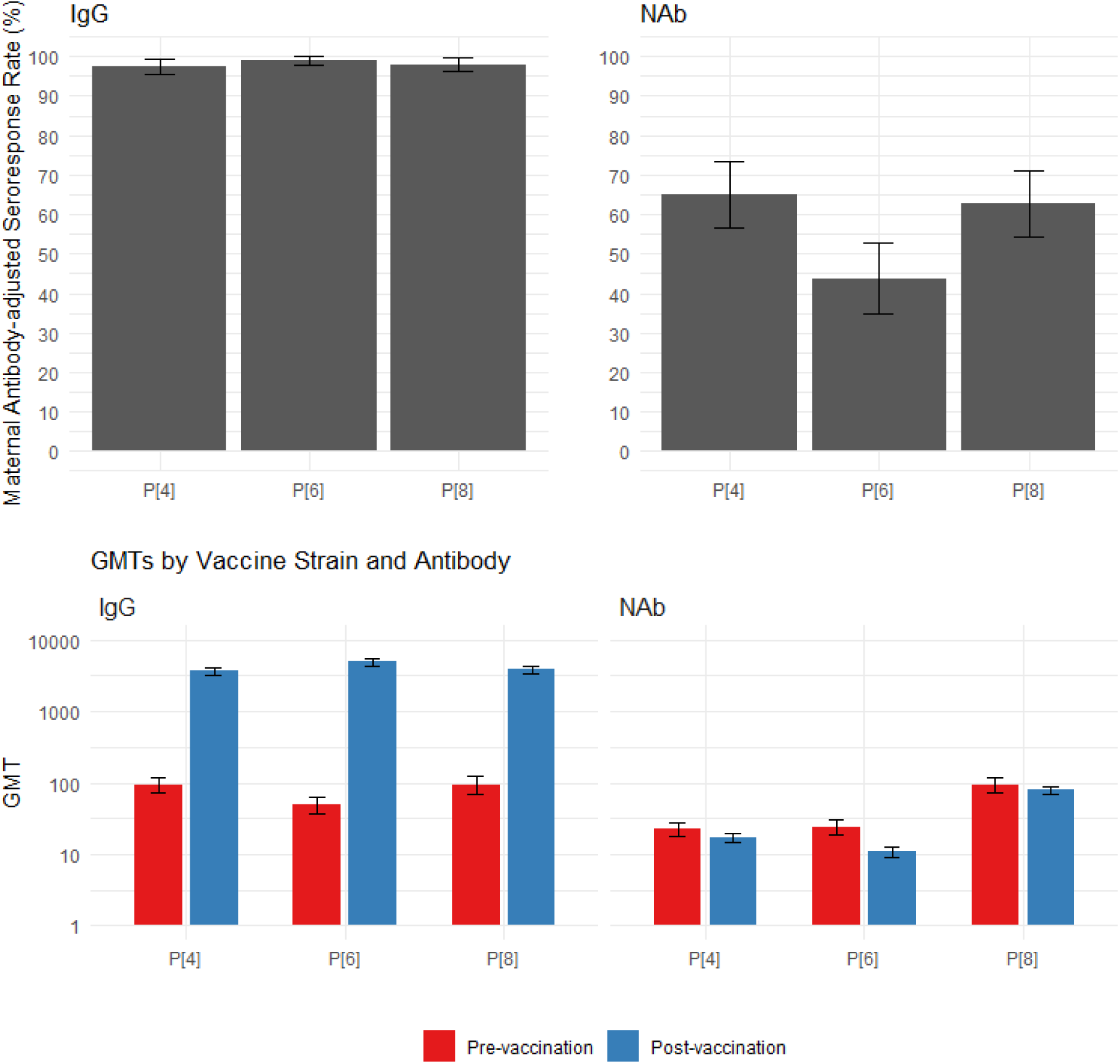
Maternal Antibody-adjusted IgG and Neutralizing Antibody (NAb) Seroresponse Rates and Geometric Mean Titers in the TV P2-VP8 arm, with 95% Confidence Intervals.

#### 4.2.3 Efficacy Results

Of 1,728 participants contributing person-time in the overall PP-EFF window in the TV P2-VP8 group, 78 reported at least one event of SRVGE ≥ 14 days after third vaccination with a crude attack rate of 4.5% (95% CI of 3.6, 5.6). Forty-five participants in the ROTARIX group reported SRVGE ≥ 14 days after third vaccination with the crude attack rate of 2.6% (95% CI of 1.9, 3.4) among 1,749 participants contributing to the person-time in the overall PP-EFF window (Table 4). The incidence rate of SRVGE ≥14 days after third vaccination per thousand person-years was 39.8 in the TV P2-VP8 group and 22.4 in the ROTARIX group. The incidence rates were higher in the TV P2-VP8 arm as compared to ROTARIX throughout the follow-up period. Cumulative incidence rates of SRVGE by time and by study arms are shown in Figure 4. The RVE in the PP-EFF population was -77.6% (95% CI of -162.3, - 21.5) (Table 4). The lower bound of the 95% CI for RVE was lower than the pre-defined non-inferiority boundary of -30% therefore, the non-inferiority of TV P2-VP8 with ROTARIX was not demonstrated. The upper confidence bound was below 0, indicating inferiority of TV P2-VP8 to ROTARIX in the prevention of SRVGE. The RVE point estimates and lower bounds of the 95% CIs were negative for all follow-up periods up to 24 months of age (Table 4; Figure 5).

**Figure 4:**
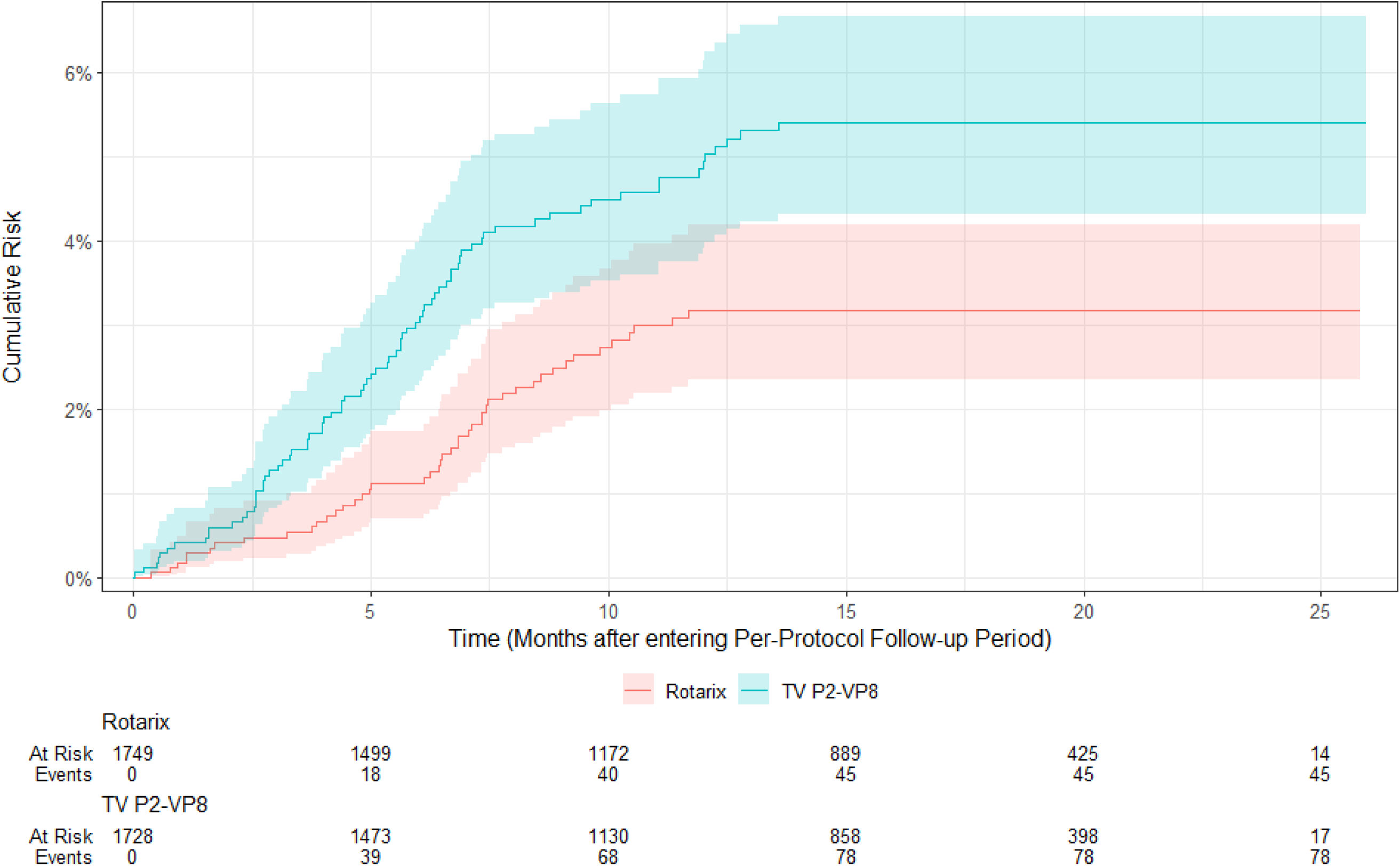
Cumulative Incidence Rate of SRVGE cases over time by Study Arms – PP-EFF Population.

**Figure 5:**
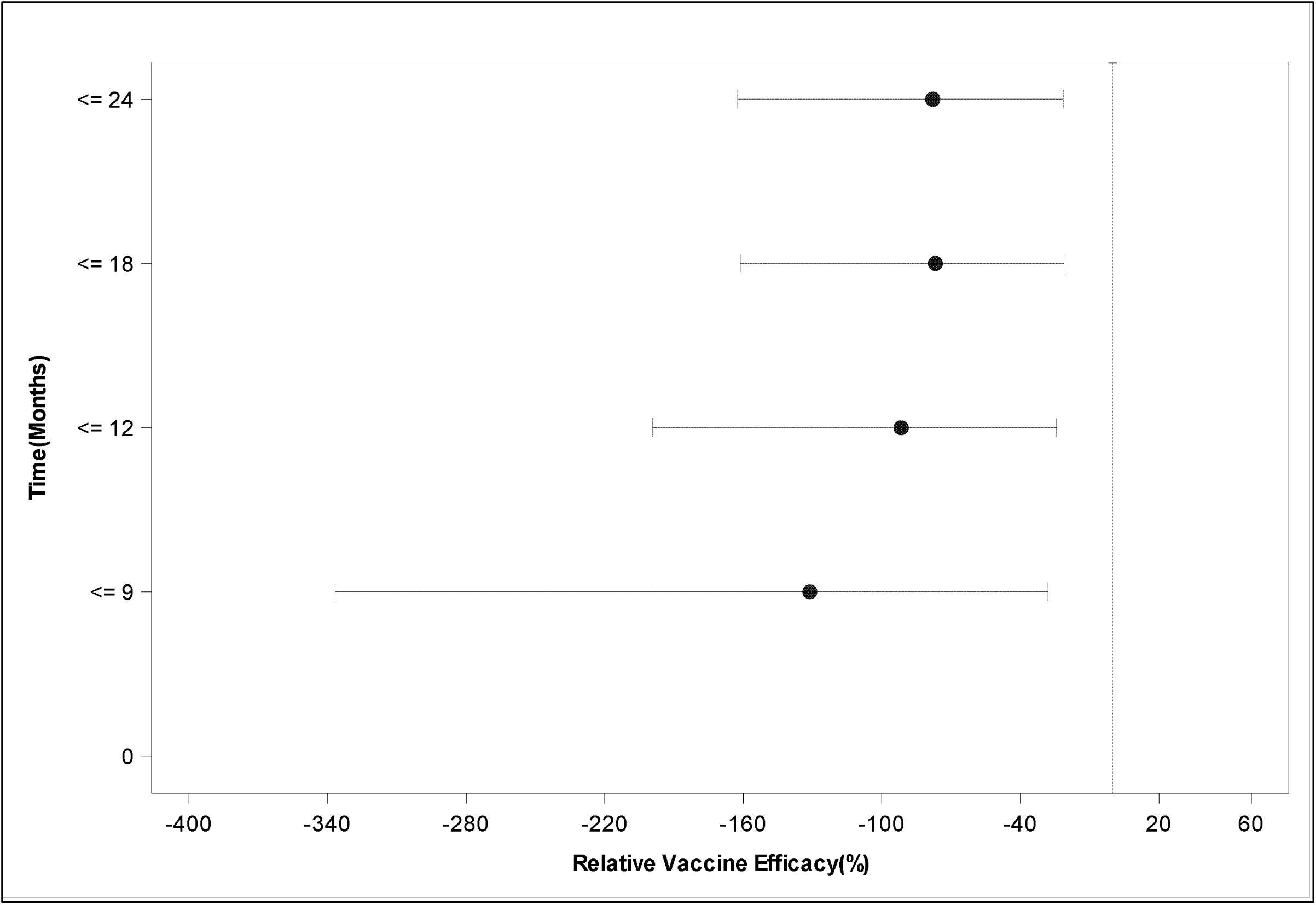
Relative Vaccine Efficacy Over Time (Months) TV P2-VP8 Arm Vs Rotarix Arm - PP-EFF Population.

**Table 4:** Incidence of Severe Rotavirus Gastroenteritis (SRVGE), Attack Rate and Relative Vaccine Efficacy – Per Protocol Efficacy Population.

| Details | Statistic | TV P2-VP8 | Rotarix | Total |
| --- | --- | --- | --- | --- |
| Number of participants contributing person-time in the PP-EFF window | N | 1728 | 1749 | 3477 |
| Number of PP-EFF participants with at least one observation of SRVGE occurring $\geq 14$ Days after Third Vaccination (E) | n | 78 | 45 | 123 |
| Crude Attack Rate (%) of SRVGE Occurring $\geq 14$ Days after Third Vaccination | $n/N \times 100$ | 4.51 | 2.57 | 3.54 |
| **Follow-up $\geq 14$ Days Post third vaccination** |  |  |  |  |
| Number of participants | n | 78 | 45 | 123 |
| Accrued follow-up time in person-years<sup>1</sup> | F | 1958.6 | 2007.1 | 3965.7 |
| Incidence rate of SRVGE Occurring $\geq 14$ Days after Third Vaccination per thousand person-years | $n/F \times 1000$ | 39.8 | 22.4 | 31.0 |
| Relative Vaccine Efficacy (RVE) | $RVE = (1-RR) \times 100$ | -77.68 | | |
| 95% CI for RVE<sup>[3]</sup> | ($LCL_{RVE}$ , $UCL_{RVE}$) | (-162.38,-21.54) | | |
| **Follow up till participant is 12 months of age (age $\leq 12$ months)** |  |  |  |  |
| Number of participants | n | 64 | 34 | 98 |
| Accrued follow-up time in person-years<sup>1</sup> | F | 1031.3 | 1048.4 | 2079.6 |
| Incidence rate of SRVGE Occurring $\geq 14$ Days after Third Vaccination per thousand person-years | $n/F \times 1000$ | 62.1 | 32.4 | 47.1 |
| Relative Vaccine Efficacy (RVE) | $RVE = (1-RR) \times 100$ | -91.67 | | |
| 95% CI for RVE<sup>[3]</sup> | ($LCL_{RVE}$ , $UCL_{RVE}$) | (-199.20,-24.35) | | |
| **Follow-up of participants between 12 to 24 months of age (both inclusive)** |  |  |  |  |
| Number of participants | n | 14 | 11 | 25 |
| Accrued follow-up time in person-years<sup>1</sup> | F | 870.5 | 900.2 | 1770.6 |
| Incidence rate of SRVGE Occurring $\geq 14$ Days after 3rd Vaccination per thousand person-years | $n/F \times 1000$ | 16.1 | 12.2 | 14.1 |
| Relative Vaccine Efficacy (RVE) | $RVE = (1 - RR) \times 100$ | -31.97 | | |
| 95% CI for RVE<sup>[3]</sup> | $(LCL_{RVE}, UCL_{RVE})$ | $(-220.37, 44.49)$ | | |

<sup>1</sup> Follow up for each participant (years) (PP efficacy analysis population) = (Minimum [Depending on participant status or date of censoring, the Date of first case of respective endpoint $\geq 14$ days post 3rd study vaccine administration, or Date of premature study discontinuation in absence of SRVGE, Date of study completion in absence of SRVGE or date of censoring due to RVGE for participants not having SRVGE] – [Date of 3rd study vaccine administration +14 (i.e. Day 15 Date)] + 1)/365.25. Accrued follow-up time in person-years = F = total follow-up time in years of all participants

The overall and site-wise crude attack rate and incidence rate of RVGE and VSRVGE in the PP-EFF population were also higher in the TV P2-VP8 group compared to the ROTARIX group, with resulting RVE being negative for all follow-up periods across sites. The frequencies of participants experiencing one or more RVGE cases [163 (9.4%) in the TV P2-VP8 group and 113 (6.5%) in the ROTARIX group] and VSRVGE cases [30 (1.7%) and 12 (0.7%) in the TV P2-VP8 and ROTARIX groups, respectively] were uniformly greater in the TV P2-VP8 group relative to the ROTARIX group (Table 5).

**Table 5:**
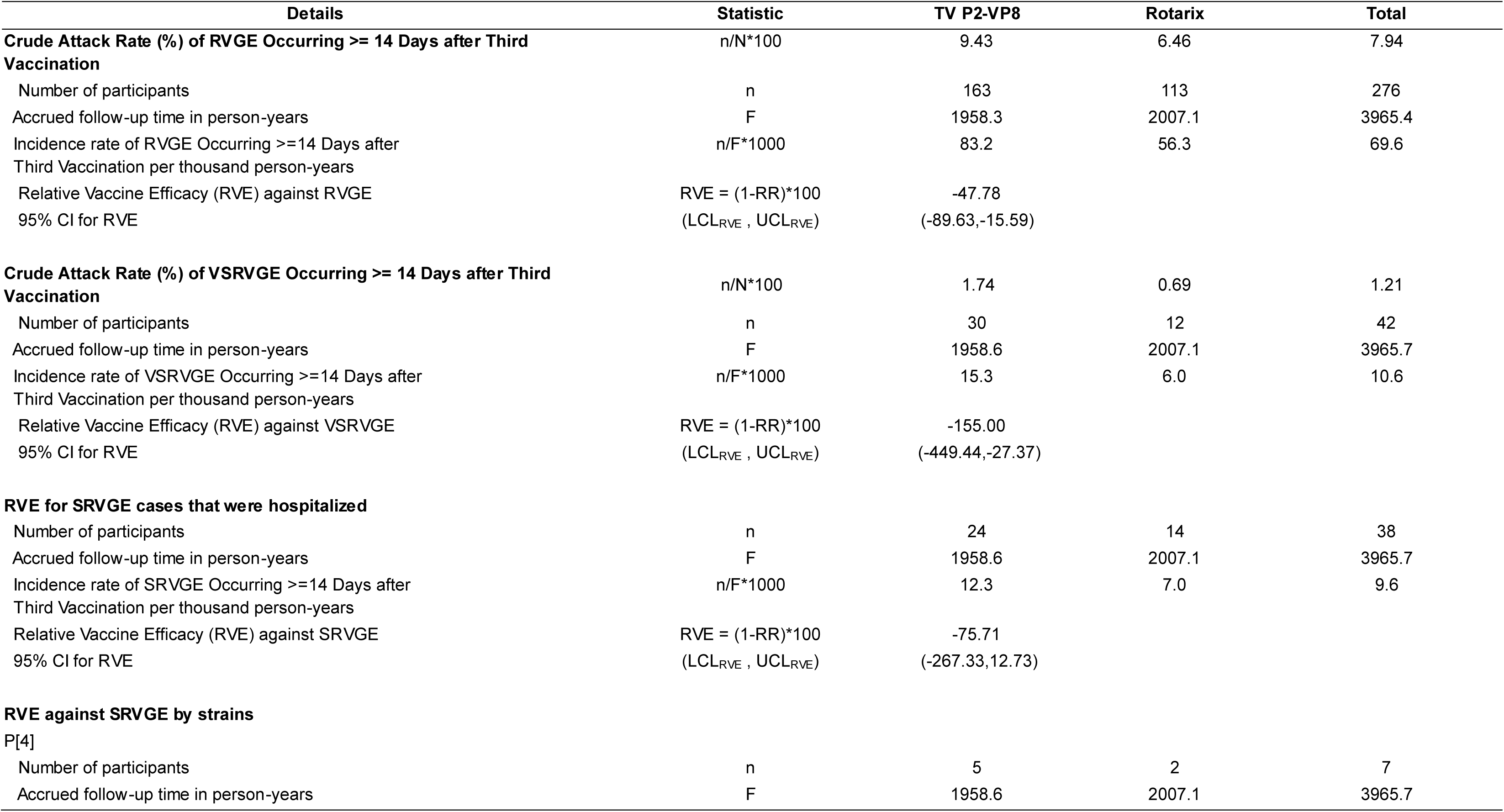
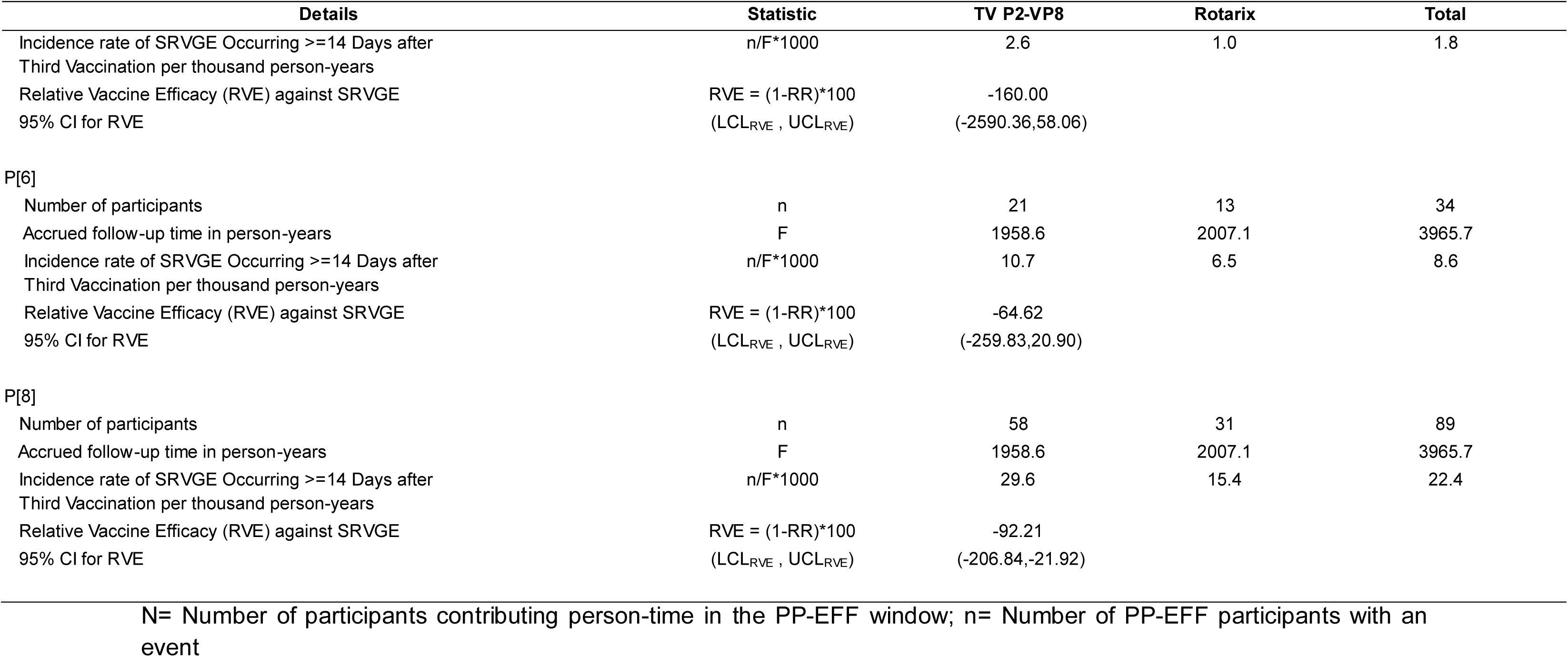
Relative vaccine efficacy against Rotavirus Gastroenteritis (RVGE), Very Severe RVGE (VSRVGE), hospitalization for Severe RVGE (SRVGE) and strain-specific SRVGE – Per-Protocol Efficacy Population.

Rotavirus strains expressing the P[8] type predominated (72%) among participants with SRVGE, followed by P[6] (27%) and P[4] (6%). The RVE against SRVGE ≥14 days after third vaccination per thousand person years for RV strains P[4], P[6], and P[8] were also negative [P[4]: -160.00 (95% CI of -2590.3, 58.6); P[6]: -64.62 (95% CI of -259.8, 20.9); P[8]: -92.21 (95% CI of -206.8, -21.9)] confirming consistency of the results for RV strains. Additionally, RVE for SRVGE cases requiring hospitalization was also negative [-75.71 (95% CI of -267.33, 12.73)] (Table 5).

A logistic regression model incorporating terms for vaccine group, site, and their interaction yielded a p-value of 0.912 for the interaction effect in the PP-EFF population. As such, RVE could not be concluded to differ across site. The ITT-EFF analysis supported the findings among the PP EFF population.

## 5. Discussion

As in previous clinical studies, the TV P2-VP8 vaccine candidate was well tolerated with no safety signals observed when concomitantly administered with routine EPI vaccines. The rate of occurrence of local and systemic solicited events was comparable between the TV P2-VP8 and ROTARIX cohorts [35.2% versus 34.1% and [74.5% versus 73.9%], respectively, with the majority of the participants presenting with Grade 1 events. The most common solicited events were injection-site pain or tenderness and fever. The fact that the local solicited events were comparable to the comparator placebo (normal saline), indicates that the local reactogenicity of TV P2-VP8 was low. Similarly, the incidence of fever was comparable in participants who received either TV P2-VP8 or ROTARIX. Fever is a commonly reported AE following the administration of the pentavalent (DPT-Hib-HepB) vaccine which was concomitantly administered with study vaccines. Serious and non-serious unsolicited AEs were similarly distributed in both the groups. The ROTARIX group reported more deaths (n=16) compared to TV P2-VP8 (n=6), though none of these were considered related to the study products.

The TV P2-VP8 vaccine candidate elicited robust serum IgG binding antibody responses (seroconversion rates >95% against all three vaccine antigens). However, the IgA binding antibody response was meager, as previously reported with this vaccine [14]. The neutralizing antibody seroconversion rate ranged from 45–65% to the three vaccine strains, which was somewhat lower than previously reported from a trial conducted in South African infants and may be study population or vaccine lot related [14]. The serum IgA response rate following immunization with ROTARIX as measured in a whole viral lysate ELISA was 46%, in line with previously published studies [6][22].

The TV P2-VP8 vaccine was inferior to ROTARIX in the prevention of SRVGE in infants (primary endpoint) as well as RVGE of any severity. RVE was consistently negative regardless of P-serotype and for each incremental 6- and 12-month age group during follow-up (<6, <12, <18, 12–24 months). Due to a lack of a placebo group, this study did not allow for determining absolute efficacy of the TV-P2-VP8 vaccine candidate. However, inversion of the RVE value (RVE of Rotarix to TV P2-VP8) yields an estimate of approximately 44% for ROTARIX, which is within the range of absolute efficacy estimates for LORVs in similar settings [23]. Therefore, protection, if any, afforded following immunization with the TV-P2-VP8 vaccine candidate is most likely minimal. Lack of discernable efficacy following immunization with TV P2-VP8 may be due to several reasons. Even though immunization with TV P2-VP8 did reduce shedding of the ROTARIX vaccine strain in a human challenge model [14], serum antibodies may only briefly translocate to the gut lumen post-immunization leading to only a transient effect. The truncated VP8 used to construct the current vaccine, being relatively small in size, may evoke a somewhat narrow immune response compared to a larger VP4 truncated protein [24]. While the vast majority of clinical isolates in this study were of the P4, P6, or P8 type, the immune response produced by the TV P2-VP8 vaccine candidate may be restricted in the epitopes recognized on the native VP4 protein expressed by rotavirus isolates currently in circulation [25]. Though serum from animals immunized with the TV P2-VP8 vaccine candidate were able to neutralize several heterologous (non-Wa) P8 strains, it is known that the VP8 region is highly variable and that only a small percentage of recently analyzed rotavirus isolates expressing the P[4], P[6], and P[8] serotype share a common lineage with the strains used to produce the vaccine [25]. A study in Bangladesh demonstrated binding antibodies against VP8 were insufficient to fully protect against SRVGE [26].

The identification of a correlate of protection is often viewed as a critical step in the development of an effective vaccine. For LORVs, serum IgA detected by ELISA using whole virus lysate as the immobilized antigen, is currently viewed as the best correlate of protection. However, this response is not viewed as mechanistic with regards to conferring immunity but rather as a marker [27]. Additionally, while useful for study populations, its predictive value does not extend to the individual level. It is unknown if this finding will extend to parenteral rotavirus vaccines, necessitating the search for preferably mechanistic correlates. Follow-up studies to identify correlates of risk are currently underway using the extensive set of serum samples collected as part of this study.

One area of focus relates to the method used in detecting neutralizing antibodies. The induction of a serum neutralizing antibody response following immunization with the TV P2-VP8 vaccine candidate did not translate into protection against rotavirus GE of any severity. This finding aligns with previous findings as immunization with LORVs induces a very modest neutralizing antibody response but still confers significant protection, questioning the protective role of such antibodies. However, a recently developed antibody-mediated intracellular neutralization (AMIN) assay may provide additional insight [28]. A study demonstrated that IgG antibodies directed against the VP6 rotavirus capsid protein were more effective at neutralizing rotavirus inside cells than VP6-specific IgA and correlated with protection in a murine rotavirus challenge model [28]. Interestingly, serum from animals immunized with the TV P-VP8 vaccine displayed a high level of activity in the conventional extracellular neutralization assay but not in the AMIN assay (unpublished).

While the early phase trials showed promise, the Phase 3 results clearly indicate that the TV P2-VP8 vaccine candidate, in its current formulation, is not effective in preventing SRVGE in this population. This study is the first to evaluate a parenteral rotavirus vaccine for efficacy against RVGE. While efficacy results were disappointing, the findings provide a basis upon which future parenteral vaccine development can be built. Future work should consider vaccines which induce a robust mucosal (IgA) response, target different or broader epitopes on rotavirus proteins, consider alternative antigens beyond VP8, and explore different delivery mechanisms that might better elicit mucosal immunity. Additional immunological analyses currently underway with serum samples obtained from participants in this study may help further elucidate immune responses potentially associated with protection from SRVGE. Scientific efforts should continue to understand the mechanism of protection afforded by LORVs and also identify alternative candidates with improved efficacy in LMICs.

## Data Availability

All the relevant data produced in the present work are contained in the manuscript. However, any additional data can be provided upon reasonable request to the corresponding author.

## Acknowledgement

The authors wish to acknowledge the support from (1) All the study participants and their parents; (2) Study staff at sites; (3) Members of the DSMB; (4) DiagnoSearch Life Sciences Pvt. Ltd. for providing project management, monitoring, medical writing, data management and statistical support;, (5) Sarah Caddy (Cornell University) for providing AMIN neutralizing antibody results.

---

**Supplementary Table 1:** Study Schema.

| **Dose** | **Study Day** | **TV P2-VP8 Arm** | **Rotarix® Arm** | **Concomitant EPI vaccines** |
| --- | --- | --- | --- | --- |
| 1 | 1 | TV P2-VP8<br>+<br>Oral Placebo (ORS) | Rotarix<br>+<br>Intramuscular Placebo<br>(normal saline) | DPT-HepB-Hib, OPV, PCV |
| 2 | 29 | TV P2-VP8<br>+<br>Oral Placebo (ORS) | Rotarix<br>+<br>Intramuscular Placebo<br>(normal saline) | DPT-HepB-Hib, OPV, PCV |
| 3 | 57 | TV P2-VP8 | Intramuscular Placebo<br>(normal saline) | DPT-HepB-Hib, OPV, PCV, IPV |
| 4/5* | - | Crossover vaccination with Rotarix<br>* | - | - |

\*Participants eligible for unblinding and cross over after interim analysis were not administered any further TV-P2 VP8, oral or IM placebo
ORS: Oral rehydration solution; DPT-HepB-Hib: Diphtheria-Tetanus-Pertussis-Hepatitis B-Haemophilus influenzae type b; OPV: Oral Polio Vaccine; PCV: Pneumococcal Conjugate Vaccine; IPV: Inactivated Poliovirus Vaccine; PCV

**Supplementary Table 2:** Overview of Reactogenicity after each dose during the 7 Days - Reactogenicity Population.

| Solicited AE | Statistic | Dose 1 |  | Dose 2 |  | Dose 3 |  |
| --- | --- | --- | --- | --- | --- | --- | --- |
|  |  | TV P2-VP8<br>(N=605) | Rotarix<br>(N=602) | TV P2-VP8<br>(N=546) | Rotarix<br>(N=528) | TV P2-VP8<br>(N=522) | Rotarix<br>(N=507) |
| **At Least one Solicited AE** | n (%), E | 354 (58.5), 917 | 360 (59.8), 849 | 279 (51.1), 669 | 265 (50.2), 597 | 272 (52.1), 580 | 252 (49.7), 544 |
| **At Least one Local Solicited AE** | n (%), E | 163 (26.9), 279 | 137 (22.8), 232 | 115 (21.1), 178 | 113 (21.4), 171 | 101 (19.3), 159 | 96 (18.9), 154 |
| Injection site pain or tenderness | n (%), E | 122 (20.2), 122 | 111 (18.4), 111 | 93 (17.0), 93 | 89 (16.9), 89 | 75 (14.4), 75 | 73 (14.4), 73 |
| Injection site erythema | n (%), E | 71 (11.7), 71 | 62 (10.3), 62 | 41 (7.5), 41 | 51 (9.7), 51 | 43 (8.2), 43 | 40 (7.9), 40 |
| Injection site induration | n (%), E | 30 (5.0), 30 | 21 (3.5), 21 | 9 (1.6), 9 | 8 (1.5), 8 | 6 (1.1), 6 | 9 (1.8), 9 |
| Injection site pruritus | n (%), E | 56 (9.3), 56 | 38 (6.3), 38 | 35 (6.4), 35 | 23 (4.4), 23 | 35 (6.7), 35 | 32 (6.3), 32 |
| **At Least one Systemic Solicited AE** | n (%), E | 333 (55.0), 638 | 342 (56.8), 617 | 253 (46.3), 491 | 242 (45.8), 426 | 246 (47.1), 421 | 232 (45.8), 390 |
| Diarrhea | n (%), E | 7 (1.2), 7 | 6 (1.0), 6 | 2 (0.4), 2 | 8 (1.5), 8 | 3 (0.6), 3 | 3 (0.6), 3 |
| Fever | n (%), E | 233 (38.5), 233 | 253 (42.0), 253 | 160 (29.3), 160 | 166 (31.4), 166 | 186 (35.6), 186 | 165 (32.5), 165 |
| Vomiting | n (%), E | 34 (5.6), 34 | 33 (5.5), 33 | 19 (3.5), 19 | 16 (3.0), 16 | 8 (1.5), 8 | 10 (2.0), 10 |
| Irritability | n (%), E | 158 (26.1), 158 | 152 (25.2), 152 | 147 (26.9), 147 | 119 (22.5), 119 | 99 (19.0), 99 | 94 (18.5), 94 |
| Decreased activity | n (%), E | 129 (21.3), 129 | 113 (18.8), 113 | 102 (18.7), 102 | 87 (16.5), 87 | 76 (14.6), 76 | 76 (15.0), 76 |
| Decreased appetite | n (%), E | 77 (12.7), 77 | 60 (10.0), 60 | 61 (11.2), 61 | 30 (5.7), 30 | 49 (9.4), 49 | 42 (8.3), 42 |

n (%), E: n = Count of Participants (at least one event i.e. Participants counted only once if the Participant reported one or more Events), % = (n / Number of Participants in Reactogenicity Population for each dose)\*100, E = Count of Events (Participant may be counted more than once)

**Supplementary Figure 1:**
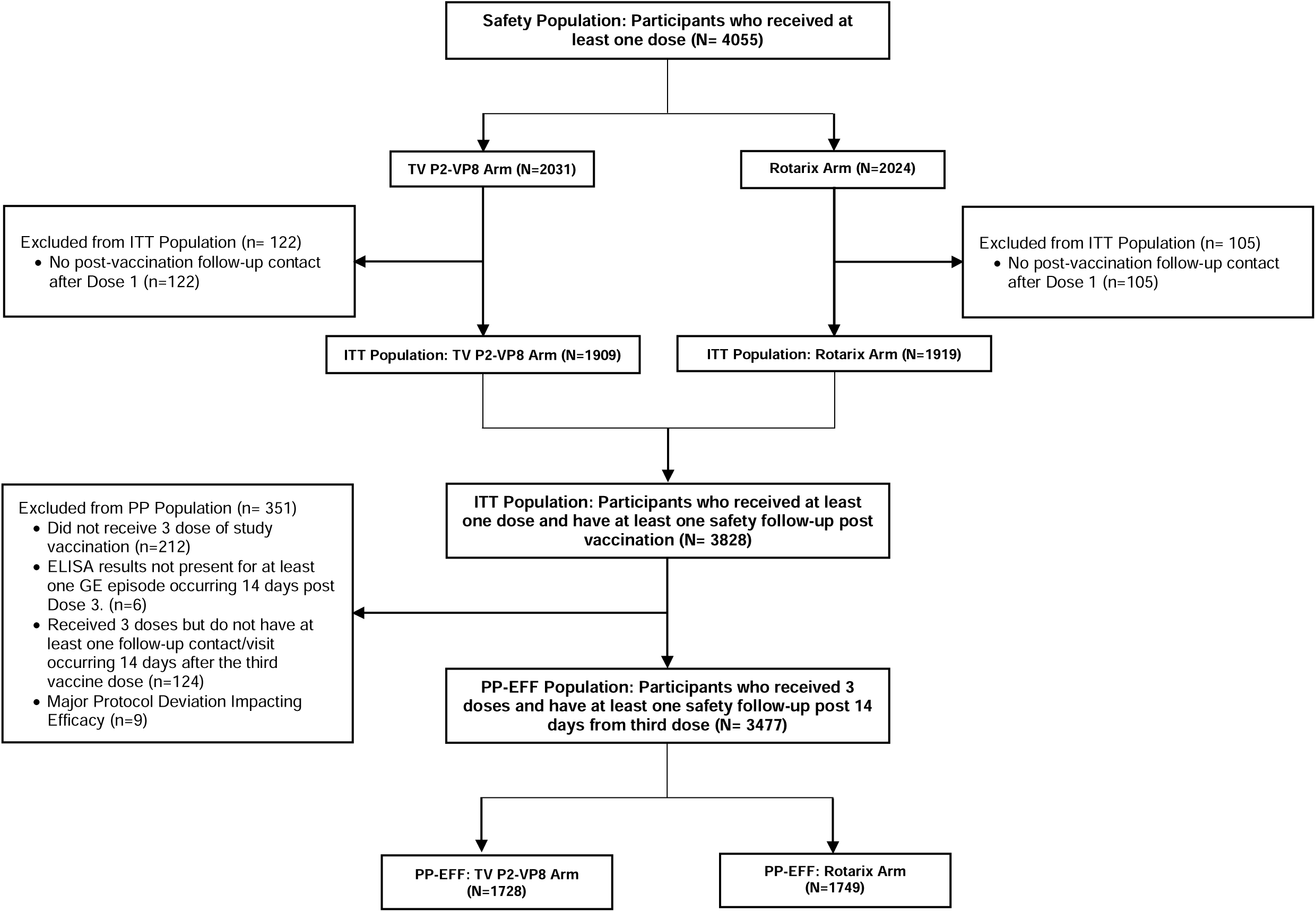
Study Populations for safety and efficacy.

## References

1. GBD 2021 Diarrhoeal Diseases Collaborators. Global, regional, and national age-sex-specific burden of diarrhoeal diseases, their risk factors, and aetiologies, 1990-2021, for 204 countries and territories: a systematic analysis for the Global Burden of Disease Study 2021. Lancet Infect Dis. 2025 May;25(5):519–536. doi: 10.1016/S1473-3099(24)00691-1. Epub 2024 Dec 18. PMID: 39708822; PMCID: PMC12018300.

2. Global immunization coverage 2024 https://www.who.int/news-room/fact-sheets/detail/immunization-coverage Accessed July 16, 2025.

3. Aliabadi N, Antoni S, Mwenda JM, Weldegebriel G, Biey JNM, Cheikh D, Fahmy K, Teleb N, Ashmony HA, Ahmed H, Daniels DS, Videbaek D, Wasley A, Singh S, de Oliveira LH, Rey-Benito G, Sanwogou NJ, Wijesinghe PR, Liyanage JBL, Nyambat B, Grabovac V, Heffelfinger JD, Fox K, Paladin FJ, Nakamura T, Agócs M, Murray J, Cherian T, Yen C, Parashar UD, Serhan F, Tate JE, Cohen AL. Global impact of rotavirus vaccine introduction on rotavirus hospitalisations among children under 5 years of age, 2008-16: findings from the Global Rotavirus Surveillance Network. Lancet Glob Health. 2019 Jul;7(7):e893–e903. doi: 10.1016/S2214-109X(19)30207-4. PMID: 31200889; PMCID: PMC7336990.

4. Vesikari T, Matson DO, Dennehy P, Van Damme P, Santosham M, Rodriguez Z, Dallas MJ, Heyse JF, Goveia MG, Black SB, Shinefield HR, Christie CD, Ylitalo S, Itzler RF, Coia ML, Onorato MT, Adeyi BA, Marshall GS, Gothefors L, Campens D, Karvonen A, Watt JP, O’Brien KL, DiNubile MJ, Clark HF, Boslego JW, Offit PA, Heaton PM; Rotavirus Efficacy and Safety Trial (REST) Study Team. Safety and efficacy of a pentavalent human-bovine (WC3) reassortant rotavirus vaccine. N Engl J Med. 2006 Jan 5;354(1):23–33. doi: 10.1056/NEJMoa052664. PMID: 16394299.

5. Vesikari T, Karvonen A, Puustinen L, Zeng SQ, Szakal ED, Delem A, De Vos B. Efficacy of RIX4414 live attenuated human rotavirus vaccine in Finnish infants. Pediatr Infect Dis J. 2004 Oct;23(10):937-43. doi: 10.1097/01.inf.0000141722.10130.50. PMID: 15602194.

6. Madhi SA, Cunliffe NA, Steele D, Witte D, Kirsten M, Louw C, Ngwira B, Victor JC, Gillard PH, Cheuvart BB, Han HH, Neuzil KM. Effect of human rotavirus vaccine on severe diarrhea in African infants. N Engl J Med. 2010 Jan 28;362(4):289–98. doi: 10.1056/NEJMoa0904797. PMID: 20107214.

7. Zaman K, Dang DA, Victor JC, Shin S, Yunus M, Dallas MJ, Podder G, Vu DT, Le TP, Luby SP, Le HT, Coia ML, Lewis K, Rivers SB, Sack DA, Schödel F, Steele AD, Neuzil KM, Ciarlet M. Efficacy of pentavalent rotavirus vaccine against severe rotavirus gastroenteritis in infants in developing countries in Asia: a randomised, double-blind, placebo-controlled trial. Lancet. 2010 Aug 21;376(9741):615-23. doi: 10.1016/S0140-6736(10)60755-6. Epub 2010 Aug 6. PMID: 20692031.

8. Bhandari N, Rongsen-Chandola T, Bavdekar A, John J, Antony K, Taneja S, Goyal N, Kawade A, Kang G, Rathore SS, Juvekar S, Muliyil J, Arya A, Shaikh H, Abraham V, Vrati S, Proschan M, Kohberger R, Thiry G, Glass R, Greenberg HB, Curlin G, Mohan K, Harshavardhan GV, Prasad S, Rao TS, Boslego J, Bhan MK; India Rotavirus Vaccine Group. Efficacy of a monovalent human-bovine (116E) rotavirus vaccine in Indian infants: a randomised, double-blind, placebo-controlled trial. Lancet. 2014 Jun 21;383(9935):2136–43. doi: 10.1016/S0140-6736(13)62630-6. Epub 2014 Mar 12. PMID: 24629994; PMCID: PMC4532697.

9. Kulkarni PS, Desai S, Tewari T, Kawade A, Goyal N, Garg BS, Kumar D, Kanungo S, Kamat V, Kang G, Bavdekar A, Babji S, Juvekar S, Manna B, Dutta S, Angurana R, Dewan D, Dharmadhikari A, Zade JK, Dhere RM, Fix A, Power M, Uprety V, Parulekar V, Cho I, Chandola TR, Kedia VK, Raut A, Flores J; SII BRV-PV author group. A randomized Phase III clinical trial to assess the efficacy of a bovine-human reassortant pentavalent rotavirus vaccine in Indian infants. Vaccine. 2017 Oct 27;35(45):6228–6237. doi: 10.1016/j.vaccine.2017.09.014. Epub 2017 Sep 26. PMID: 28967523; PMCID: PMC5651219.

10. Neuzil KM, Parashar UD, Steele AD. Rotavirus vaccines for children in developing countries: understanding the science, maximizing the impact, and sustaining the effort. Vaccine 2012;30 Suppl 1:A1–2.

11. Patel M, Shane AL, Parashar UD, Jiang B, Gentsch JR, Glass RI. Oral rotavirus vaccines: how well will they work where they are needed most? Journal of Infectious Diseases 2009;200 Suppl 1:S39–48.

12. Fix AD, Harro C, McNeal M, Dally L, Flores J, Robertson G, Boslego JW, Cryz S. Safety and immunogenicity of a parenterally administered rotavirus VP8 subunit vaccine in healthy adults. Vaccine. 2015 Jul 17;33(31):3766–72. doi: 10.1016/j.vaccine.2015.05.024. Epub 2015 Jun 8. PMID: 26065919.

13. Groome MJ, Koen A, Fix A, Page N, Jose L, Madhi SA, McNeal M, Dally L, Cho I, Power M, Flores J, Cryz S. Safety and immunogenicity of a parenteral P2-VP8-P[8] subunit rotavirus vaccine in toddlers and infants in South Africa: a randomised, double-blind, placebo-controlled trial. Lancet Infect Dis. 2017 Aug;17(8):843–853. doi: 10.1016/S1473-3099(17)30242-6. Epub 2017 May 5. PMID: 28483414; PMCID: PMC7771518.

14. Groome MJ, Fairlie L, Morrison J, Fix A, Koen A, Masenya M, Jose L, Madhi SA, Page N, McNeal M, Dally L, Cho I, Power M, Flores J, Cryz S. Safety and immunogenicity of a parenteral trivalent P2-VP8 subunit rotavirus vaccine: a multisite, randomised, double-blind, placebo-controlled trial. Lancet Infect Dis. 2020 Jul;20(7):851–863. doi: 10.1016/S1473-3099(20)30001-3. Epub 2020 Apr 3. PMID: 32251641; PMCID: PMC7322558.

15. Ruuska T, Vesikari T. Rotavirus disease in Finnish children: use of numerical scores for clinical severity of diarrhoeal episodes. Scand J Infect Dis. 1990;22(3):259–67. doi: 10.3109/00365549009027046. PMID: 2371542.

16. Premier® Rotaclone® package insert https://www.meridianbioscience.com/uploads/696004_pi.pdf?country=IN Accessed July 16, 2025

17. Division of AIDS (DAIDS) Table for Grading the Severity of Adult and Pediatric Adverse Events. https://rsc.niaid.nih.gov/sites/default/files/daidsgradingcorrectedv21.pdf Accessed Aug 12, 2025

18. Knowlton DR, Spector DM, Ward RL. Development of an improved method for measuring neutralizing antibody to rotavirus. J Virol Methods. 1991 Jun;33(1-2):127–34. doi: 10.1016/0166-0934(91)90013-p. PMID: 1658027.

19. Jennison C, Turnbull BW. Group sequential methods with applications to clinical trials. CRC Press; 1999 Sep 15.

20. Anderson K (2025). _gsDesign: Group Sequential Design_. R package version 3.6.7, https://CRAN.R-project.org/package=gsDesign

21. R Core Team (2024). _R: A Language and Environment for Statistical Computing_.R Foundation for Statistical Computing, Vienna, Austria. https://www.R-project.org/

22. Madhi SA, Kirsten M, Louw C, Bos P, Aspinall S, Bouckenooghe A, Neuzil KM, Steele AD. Efficacy and immunogenicity of two or three dose rotavirus-vaccine regimen in South African children over two consecutive rotavirus-seasons: a randomized, double-blind, placebo-controlled trial. Vaccine. 2012 Apr 27;30 Suppl 1:A44–51. doi: 10.1016/j.vaccine.2011.08.080. PMID: 22520136.

23. Henschke N, Bergman H, Hungerford D, Cunliffe NA, Grais RF, Kang G, Parashar UD, Wang SA, Neuzil KM. The efficacy and safety of rotavirus vaccines in countries in Africa and Asia with high child mortality. Vaccine. 2022 Mar 15;40(12):1707–1711. doi: 10.1016/j.vaccine.2022.02.003. Epub 2022 Feb 17. PMID: 35184924; PMCID: PMC8914343.

24. Chen Y, Luo G, Song F, Wang X, Zhang S, Ge S, Li T, Zhang J, Xia N. Truncated rotavirus VP4 proteins induce stronger protective immunity compared to P2 - VP8 in animal models. Antiviral Res. 2025 Jun;238:106156. doi: 10.1016/j.antiviral.2025.106156. Epub 2025 Apr 5. PMID: 40194664.

25. Velasquez DE, Jiang B. Evolution of P[8], P[4], and P[6] VP8* genes of human rotaviruses globally reported during 1974 and 2017: possible implications for rotavirus vaccines in development. Hum Vaccin Immunother. 2019;15(12):3003–3008. doi: 10.1080/21645515.2019.1619400. Epub 2019 Jun 13. PMID: 31124743; PMCID: PMC6930099.

26. Lee B, Colgate ER, Carmolli M, Dickson DM, Gullickson S, Diehl SA, Ara R, Alam M, Kibria G, Abdul Kader M, Afreen S, Ferdous T, Haque R, Kirkpatrick BD. Plasma VP8∗-Binding Antibodies in Rotavirus Infection and Oral Vaccination in Young Bangladeshi Children. J Pediatric Infect Dis Soc. 2022 Apr 30;11(4):127–133. doi: 10.1093/jpids/piab120. PMID: 34904667; PMCID: PMC9055852.

27. Angel J, Steele AD, Franco MA. Correlates of protection for rotavirus vaccines: Possible alternative trial endpoints, opportunities, and challenges. Hum Vaccin Immunother. 2014;10(12):3659–71. doi: 10.4161/hv.34361. PMID: 25483685; PMCID: PMC4514048.

28. Caddy SL, Vaysburd M, Wing M, Foss S, Andersen JT, O’Connell K, Mayes K, Higginson K, Iturriza-Gómara M, Desselberger U, James LC. Intracellular neutralisation of rotavirus by VP6-specific IgG. PLoS Pathog. 2020 Aug 4;16(8):e1008732. doi: 10.1371/journal.ppat.1008732. PMID: 32750093; PMCID: PMC7428215.

